# A comparative study of SIR Model, Linear Regression, Logistic Function and ARIMA Model for forecasting COVID-19 cases

**DOI:** 10.1101/2021.05.24.21257594

**Authors:** Saina Abolmaali

## Abstract

Starting February 2020, COVID-19 was confirmed in 11,946 people worldwide, with a mortality rate of almost 2%. A significant number of epidemic diseases including human Coronavirus display patterns. In this study with the benefit of data analytic, we develop regression models and a Susceptible-Infected-Recovered (SIR) model for the contagion to compare the performance of models to predict number of cases. first, we implement a good understanding of data and perform Exploratory Data Analysis (EDA). Then, we derive the parameters of the model from the available data corresponding to the top 4 regions based on the history of infections and the most infected people as of the end of August 2020. Then models are compared and further research are introduced.

## 1 Introduction

A pandemic is defined as “an epidemic occurring world-wide, over a very wide area, crossing international boundaries, and usually affecting a large number of people”[1]. Since this is a broad definition that could include seasonal epidemics (which are not considered pandemics), the transmissibility and severity of a disease can be measured to characterize and further describe it. One metric used to measure the transmissibility of a disease is the effective reproduction number (R), which represents the average number of persons infected by one single infectious individual. A measure of severity is the case fatality ratio, which represents the number of deaths caused by the disease. The World Health Organization (WHO) lists nineteen (19) pandemic, epidemic diseases: Chikungunya, Cholera, Crimean-Congo haemorrhagic fever, Ebola virus, Hendra virus infection, Influenza (pandemic, seasonal, zoonotic), Lassa fever, Marburg virus disease, Meningitis, MERS-CoV, Monkeypox, Nipah virus infection, Plague, Rift Valley fever, SARS, Smallpox, Tularaemia, Yellow fever, and Zika virus disease[2]. On March 11, 2020, the WHO declared the novel coronavirus (2019-nCoV) a global pandemic, adding a twentieth disease to this list[3]. On April 25, 2020, the number of confirmed cases reached 2,810,325 and the number of confirmed deaths 193,825, affecting in this way 213 countries, areas, or territories[4]. Although there are still many questions about this disease, data is being collected and used to learn more about this disease. This study seeks to predict the number of confirmed cases and the number of deaths with the epidemic model and data analytical models. Data analytic have been used in many different areas such as transportation, finance[5] and healthcare. Pandemics have been a topic of interest to several researchers in the data analytic field. Consequently, researchers have been used different models to study the behavior of the data, gain some insight, and draw conclusions. One popular model that is being used is the SIR model. One of the most recent pandemics (before COVID-19) was the H1N1[6]. According to the Centers for Disease Control and Prevention (CDC), between April 12, 2009 and April 10, 2010 the number of cases reported were 60.8 million and the number of deaths 12,469 in the United States[7]. Ebola (first discovered in 1976) had a recent large outbreak in West Africa (2014-2016). In this significant outbreak, there were 28,652 cases and 11,325 deaths according to the CDC[8]. Chowell et al.[9] discussed the most common modelling approaches used to study and analyze the early spread of an epidemic. These approaches include meta population spatial models, individual-based network models, examining early growth from spatial models (which includes the SIR model), SIR model with reactive behavior changes, and SIR model with in homogeneous mixing. The authors identified a gap that requires the incorporation of important epidemic features, such as a flexible epidemic growth (from polynomial to exponential dynamics). Mutalik [10] provided a literature review of mathematical models used to predict H1N1 outbreaks. The author included thirty-one (31) articles; nine (9) of them used SIR model and the other nine (9) us SIER model. Other models included: SIS; Compartmental Model; combined model; combined model with SIER – two models only; early exponential growth rate, simple SIER model and complex SIER model, stochastic SIR model; combination of SIS, SIR, SIER. The author found that the most used mathematical model was the SIER model. The author concluded that a mathematical model along with another secondary model will generate a better prediction. Zhan et al.[11] used COVID-19 historical data of 367 cities in China and obtained the set of parameters of the augmented Susceptible - Exposed-Infected-Removed (SEIR) model for each city, to create a set of profile codes representing a variety of transmission mechanisms and contact topology. They compared the data of an early outbreak of a given population with the complete set of historical profiles. Then, they selected the best fit profiles and used the corresponding sets of profile codes for prediction of the future progression of the epidemic in that population. They applied the method to the data of South Korea, Italy and Iran. The results showed that peaks of infection cases were expected to happen before the end of March 2020. Moreover, the percentage of population infected in each city would be less than 0.01%, 0.05% and 0.02%, for South Korea, Italy and Iran, respectively. In another research Lover and McAndrew[12] used exponential growth model and epidemiological parameters from the epidemic in Wuhan, China to forecast cumulative infections in the United States. Their forecast results showed that a significant number of infections are undetected, and without considerable non-pharmaceutical interventions, the number of infections should be expected to grow exponentially. In another work Liu et al.[13] used SEIR model combined with network-driven dynamics to simulate the spread of COVID-19 in the United States accounting for the domestic air traffic occurring amongst the 50 US states, Washington DC, and Puerto Rico. Based on the model predictions for March 14 to March 16, if no containment plans were done, the national epidemic peak could be expected to arrive by early June, corresponding to a daily active count of 7% of the US population. Their results showed that Epidemic peaks were expected to arrive in the Washington and New York states by May 21 and 25, respectively. They also reported that the epidemic progression could be delayed by up to 34 days with a modest 25% reduction in COVID-19 transmissibility via community-level interventions. Another model was implemented by Roosa et al.[14]. They used three phenomenological models to do short-term forecasts in real-time. The models had been previously used to perform short-term forecasts for several infectious diseases, including SARS, Ebola, pandemic influenza, and dengue. The generalized logistic growth model (GLM) extended the simple logistic growth model to accommodate sub-exponential growth dynamics with a scaling of growth parameter, p. The Richards model also included a scaling parameter, a, to allow for deviation from the symmetric logistic curve. They also included a sub-epidemic wave model that supports complex epidemic trajectories, including multiple peaks. Based on data up until February 9, 2020, their forecasts agreed across the three models presented to a large extent and predicted an average range of 7409–7496 additional confirmed cases in Hubei and 1128–1929 additional cases in other provinces within the next five days. Models also predicted an average total cumulative case count between 37,415 and 38,028 in Hubei and 11,588–13,499 in other provinces by February 24, 2020. Taking into account the nature of the epidemic disease data is time series, Gupta and Pal [15] applied ARIMA model to predict the future trends in India. Based on their forecasts generated by ARIMA model, number of infected cases in India may go up to 700 thousands in the next 30 days in worst case scenario. However, most optimistic scenario may show the numbers up to 1000-1200. Moreover, the average number of infected cases predicted by ARIMA model was around 7000 in next 30 days while the current numbers was 536. Anastassopoulou et al. [16] used the Susceptible-Infectious-Recovered-Dead (SIDR) model and data of the COVID-19 spread in Hubei, China from January 11 to February 10, 2020 to estimate the parameters of basic reproduction number *R*_0_ (2.6 based on confirmed cases and almost 2 considering twenty times the number of confirmed cases and forty times the number of recovered) and per day infection mortality (0.15% considering the second scenario) and recovery rates. The authors also predicted that the epicenter would be on February 29, 2020 with a cumulative number of infected of 45,000-180,000 and number of deaths of more than 2,700. Read et al.[17] applied a fitted a deterministic SEIR meta population transmission model with an assumed four (4) days incubation period (based on a SARS approximation) to estimate the *R*_0_, which ranges between 3.6 and 4.0. Moreover, they estimated a transmission rate of 1.07 within Wuhan. The authors estimate that only 5.1% (with a 95% confidence interval) of infections in Wuhan are identified. They also predict more than 190,000 cases by February 4, 2020. Riou and Althaus [13] simulated early outbreak trajectories and estimated the *R*_0_ to be around 2.2 (in a 90% interval, 1.4–3.8). The authors compared the early pattern of human-to-human transmission of COVID-19 to past viruses and concluded that is similar to SARS-CoV (2002).

## 2 Method

### 2.1 Data

For this research we have used GitHub data repository managed by Johns Hopkins University which contains daily time series summary tables, including confirmed, deaths and cases infected for more than once per day. Daily data of the influenced individuals are very helpful for data scientists. All data are from the daily case report, retrieved from: https://github.com/CSSEGISandData/COVID-19. We applied three informational data sets in our models. The main informational collection is the confirmed cases data set comprises every day information of the number of confirmed cases. The subsequent data set is the death data set that has every day data of the number of dead individuals. The most important one is the last data set including the recovered ones, which gives the absolute data on the quantity of recovered cases in the data collection. The data has been accessible since January 22th of 2020.

### 2.2 EDA Analysis

The number of global confirmed cases and deaths since January 22 are graphically illustrated in Figure 1. It shows that the expansion begins between March and April 2020. It is important to note that this number includes the reported cases of people who have been tested. No nation knows the actual number of individuals tainted with COVID-19. All we know is the status of the individuals who have been tested. Each of them who has a lab-affirmed contamination is considered as a confirmed case. This implies the tallies of affirmed cases rely upon how much a nation really tests and the reliability of results correctness. To decipher any information on the confirmed cases we have to know how many testing for COVID-19 the nation really does. Although these numbers do not exactly reflect the real situation that world is facing, they still give a valuable insight about the behavior of this disease growth. We broke down our data sets with various EDA techniques and envisioned those information to give an adequate cognizance with respect to the flare-up of COVID-19. Top four nations with the most infected patients are USA, Brazil, India, Russia, South Africa and Peru. We have separated the quantity of confirmed cases and the fatalities to show how Coronavirus is contaminating individuals in each country. This measurement offers two key experiences: initially as a proportion of how sufficient nations are testing; and furthermore to assist us with understanding the spread of the infection, related to information on confirmed cases. The positive rate is a decent measurement for how satisfactorily nations are trying in light of the fact that it shows the degree of testing compared to the size of the episode. To have the option to appropriately screen and control the spread of the infection, nations with more boundless flare-ups need to accomplish all the more testing. For classification, regression or forecast of a specific issue, feature selection techniques can be utilized to discover the highlights that have the most elevated effect on that issue.

**Fig. 1.**
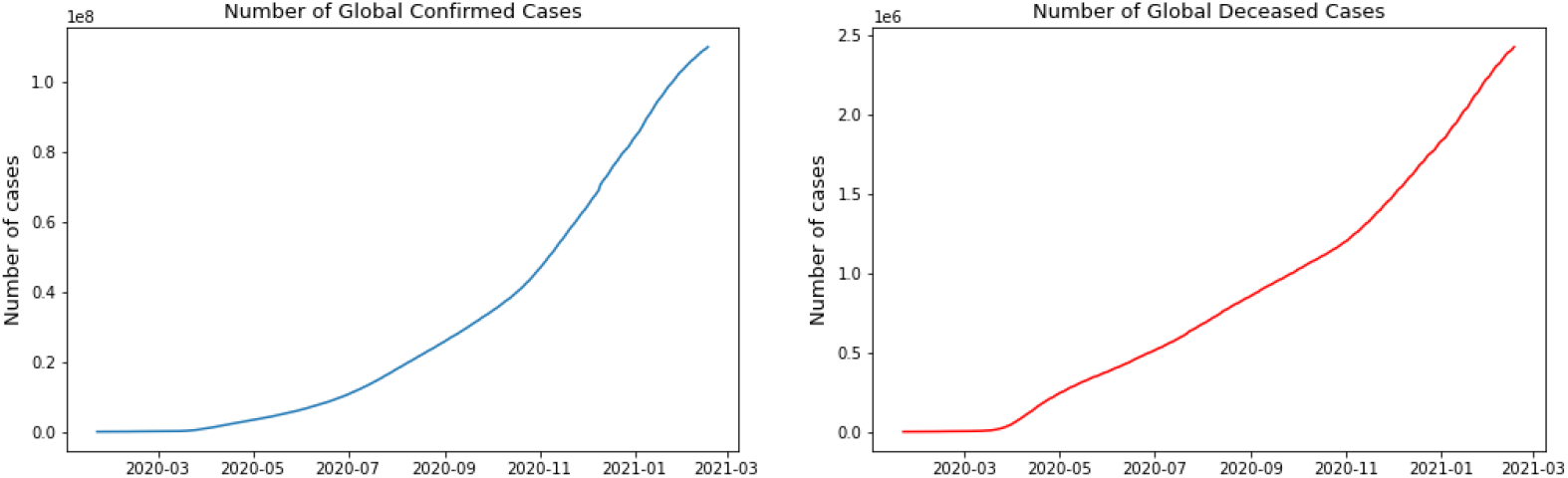
Global number of confirmed cases and deceased cases

As indicated by Figure 2, it doesn’t appear that the spread is controlled in any of the referenced nations. As we can see in the depicted charts, USA has the highest number of infected patients. The figure shows that in almost 4 month of the first case announced in United State more than 5 million people were infected and after that India has the sharpest rate of infection.

**Fig. 2.**
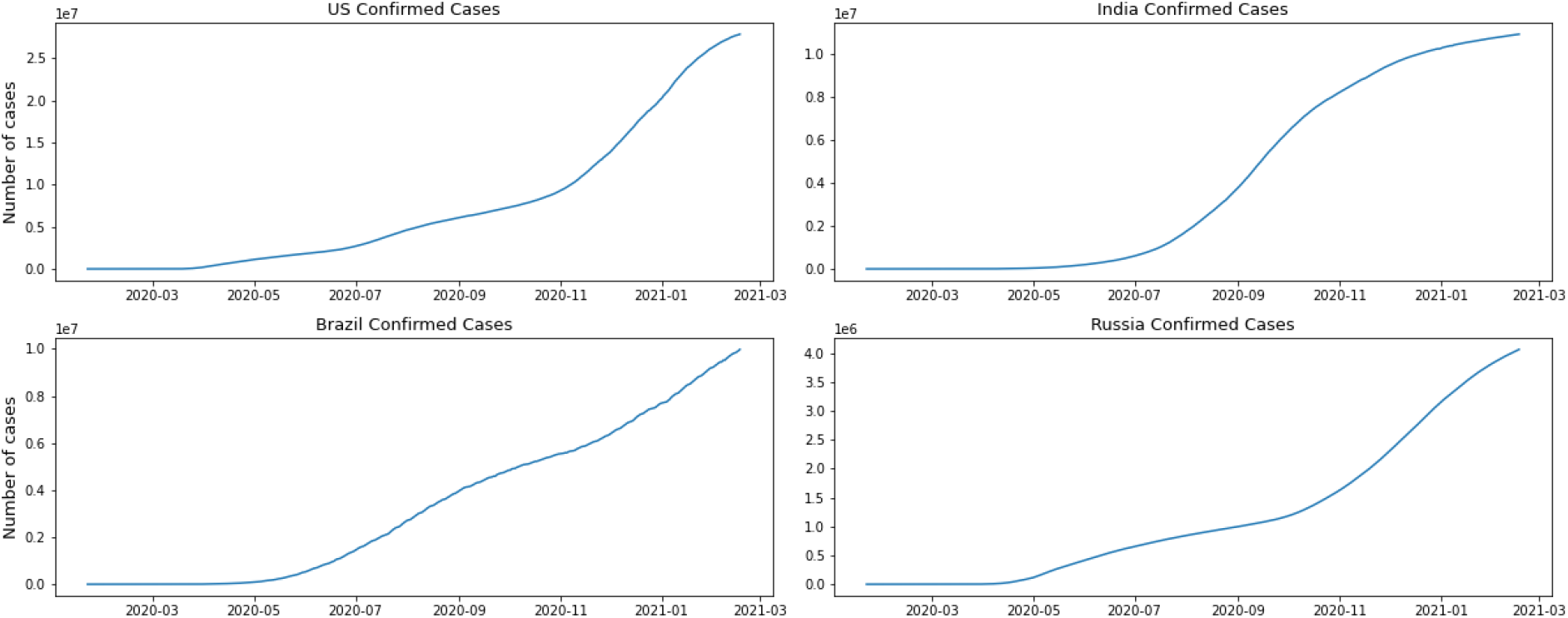
Confirmed cases per country

### 2.3 SIR Model

The mathematical modelling of epidemics has been the object of a vast number of studies over the past century[18]. Given the importance of epidemics for life on Earth in general, it is not in the least astonishing that the desire to understand their mechanism has led to the formulation of models which make possible the simulation of events for which laboratory experiments cannot be conducted easily [19]. The reason we have chosen SIR model is that there is not enough evidence that the patient might not be immune to the disease. Prominent among the mathematical models of epidemics, and of great historical importance, is the susceptible– infected-removed (SIR) model initially proposed by Kermack and McKendrick[20]. The model has been defined with three groups of healthy people who are susceptible (S), infected individuals (I), removed individuals either by them being recovered and immunized or by their death (R). Since the number of susceptible, infected and recovered people may fluctuate over time, SIR model is a dynamic model. Flowing from susceptible to infected and then recovered could be showed in the figure 4

In this model the infection rate is *β*, which is the probability transmitting disease between a susceptible and an infectious individual. *γ* is the recovery rate. *N* is defined as population and is equal to *N* = *S* + *I* + *R*. We can write the SIR model as the following differential equation:

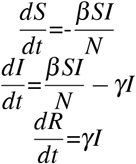

To perform the SIR model we have started with 1000 as the number of population. We have used initial number of infected equal to one and initial number of removed equal to zero as the data set.Therefore, everyone else is susceptible to infection initially.^1^. After taking several tests on the model we have observed that the best combination of the beta and gamma for our data set would be *β* = 3.524 which is a mean number of contacts (sufficient to spread the disease) per day that each infected individual has) and also *γ* = 3.45 the infected group that recovers(or dies) during any given day. This combination will give us the SIR model as show in the following figure. In this model we did not consider the influence of immigration because once an epidemic has started, the influence of any additional immigrants is small. In fact, the relative influence of an immigrant in the subsequent growth of the epidemic drops geometrically with the number of local infected[22]. After calculating the best fit parameters of the model we have run the model and we have for our data set on the model. The following figure shows the best possible fit of the data for US. The model does not show a good fit for the number of the infected individuals. The graphical result shows that the SIR Model can not provide useful early prediction of the epidemic in this case. To improve we have decided to move to regression analysis.

### 2.4 Linear Regression

Our general surroundings is profoundly muddled. For instance, how an infection spreads, including the novel strand of Coronavirus (SARS-CoV-2) that was distinguished in Wuhan, China, relies on numerous components, among which some of them are considered by the exemplary SIR model, which is somewhat over-simplified and can’t contemplate floods in the quantity of susceptible people. Regression models are utilized to assess or anticipate the target variable based on dependent factors. As we know regression modeling characterizes an influential technique to model and estimate the target variable. For the instance of predicting a continuous amount response variable regression is utilized, while classification is reasonable for foreseeing a discrete class label response. Subsequently for demonstrating the quantity of confirmed cases after some time and anticipating the future development, regression is thought of. To model the relationship between the response and the explanatory variable we are going to use linear regression. Therefore we can see in the plots, in some cases the linear regression can define the Covid data. Although the linear regression can define the Covid, we cannot use it to prediction in long term. We have used the data to try to predict in long term, the results are not satisfying.

**Fig. 3.**
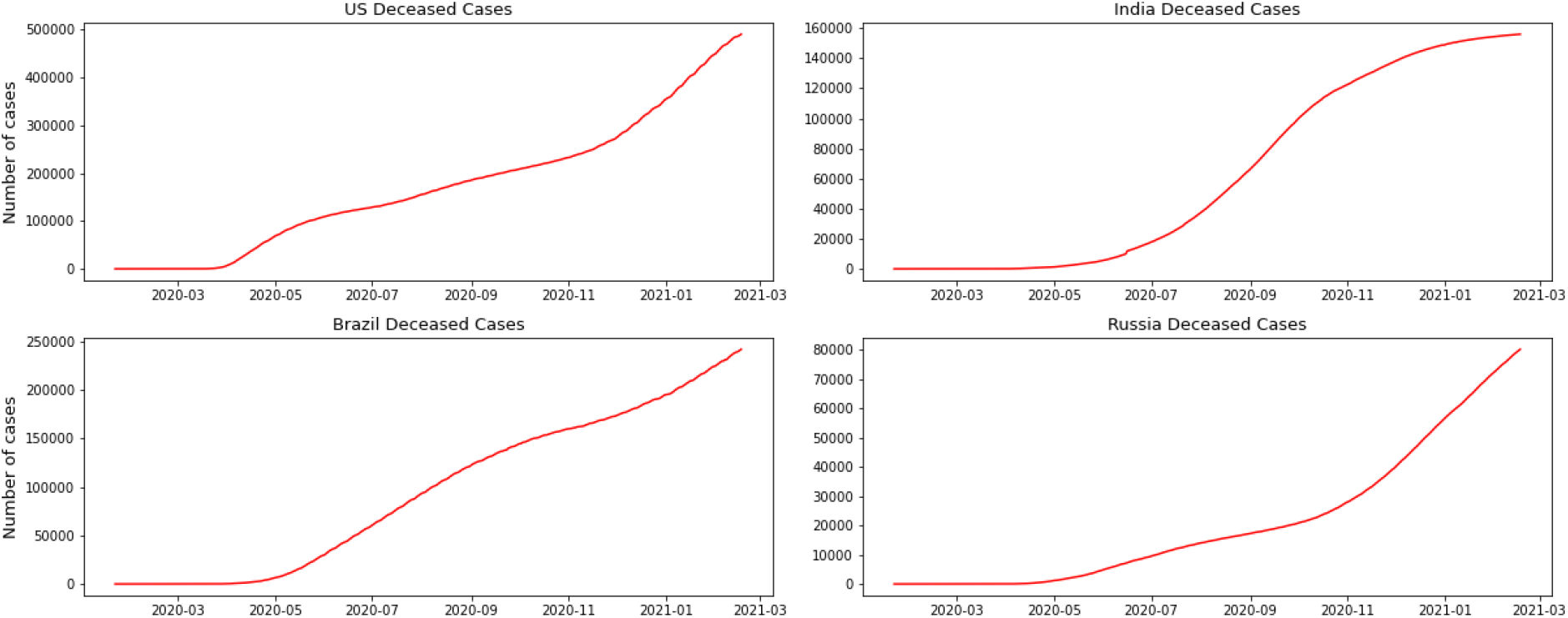
Deceased cases per country

**Fig. 4.**
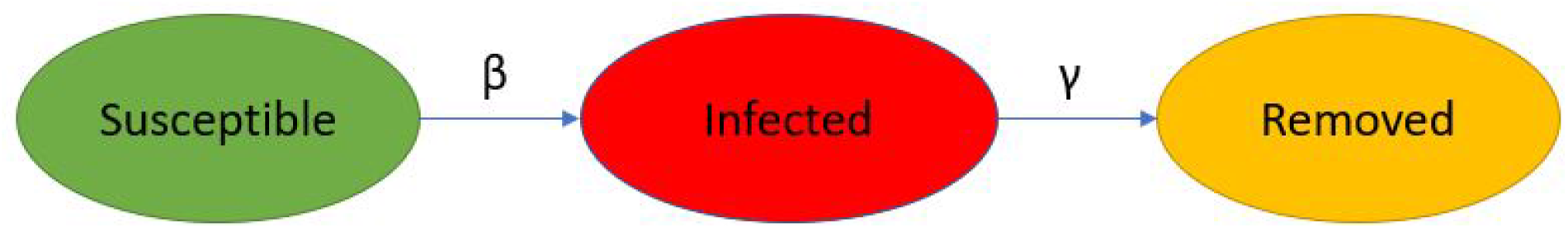
SIR Following

### 2.5 Logistic Function

logistic equation was initially advanced in 1920 not as an advantageous depiction, yet as a law of development, and was enthusiastically condemned by statisticians and biologists for the resulting decade and a half. However it endured and rose in an alternate setting as one of the base models of experimental populace biology in the 1930’s and 1940’s. The move from dismissal to acknowledgment was in no way, easy and was not just because of biologists’ progressive acknowledgment of the natural value of the curve. The logistic curve portrays the development of a populace after some time. In its easiest structure it is S-shaped, balanced, and is portrayed with the equation:

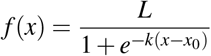

**Fig. 5.**
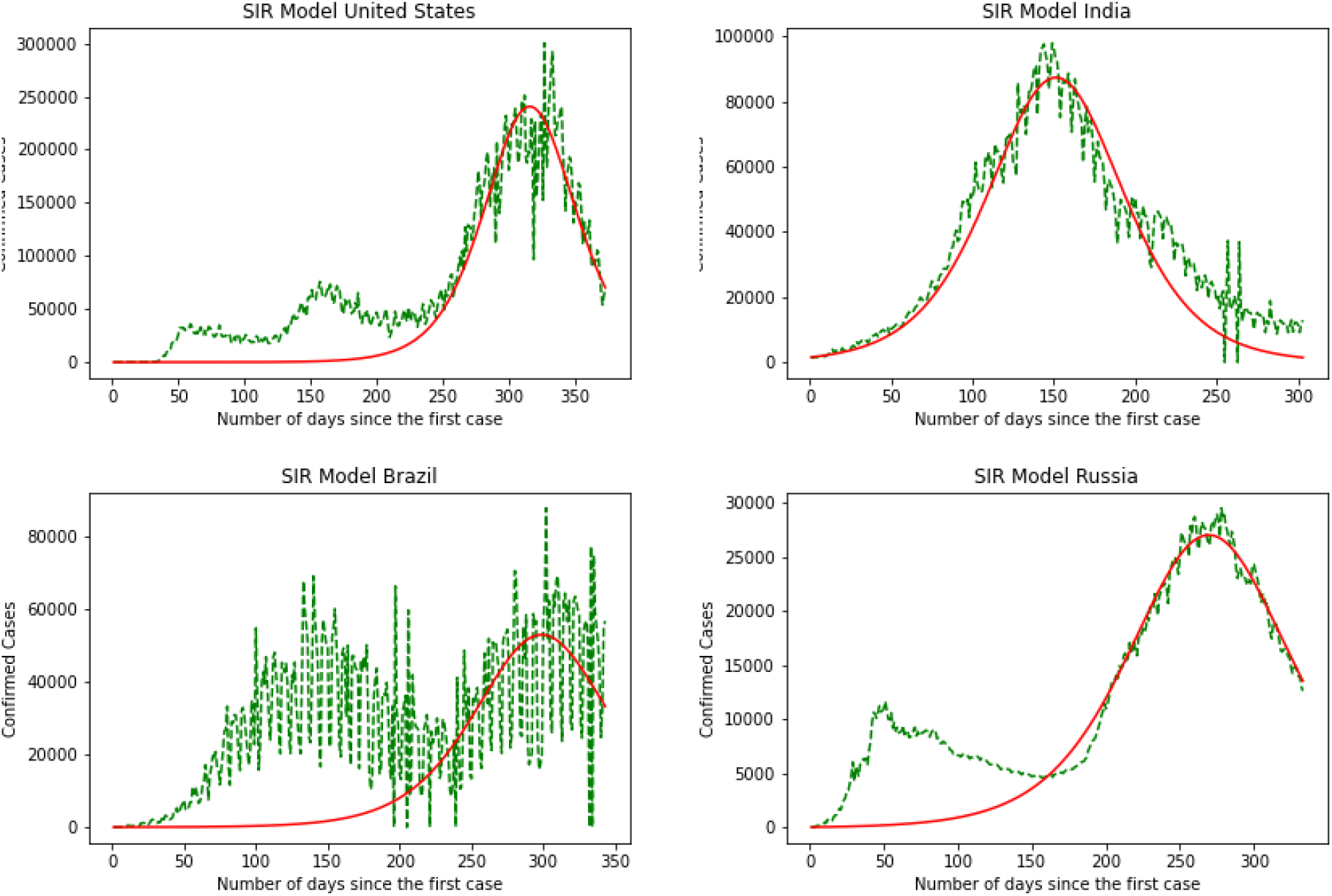
SIR Model

**Fig. 6.**
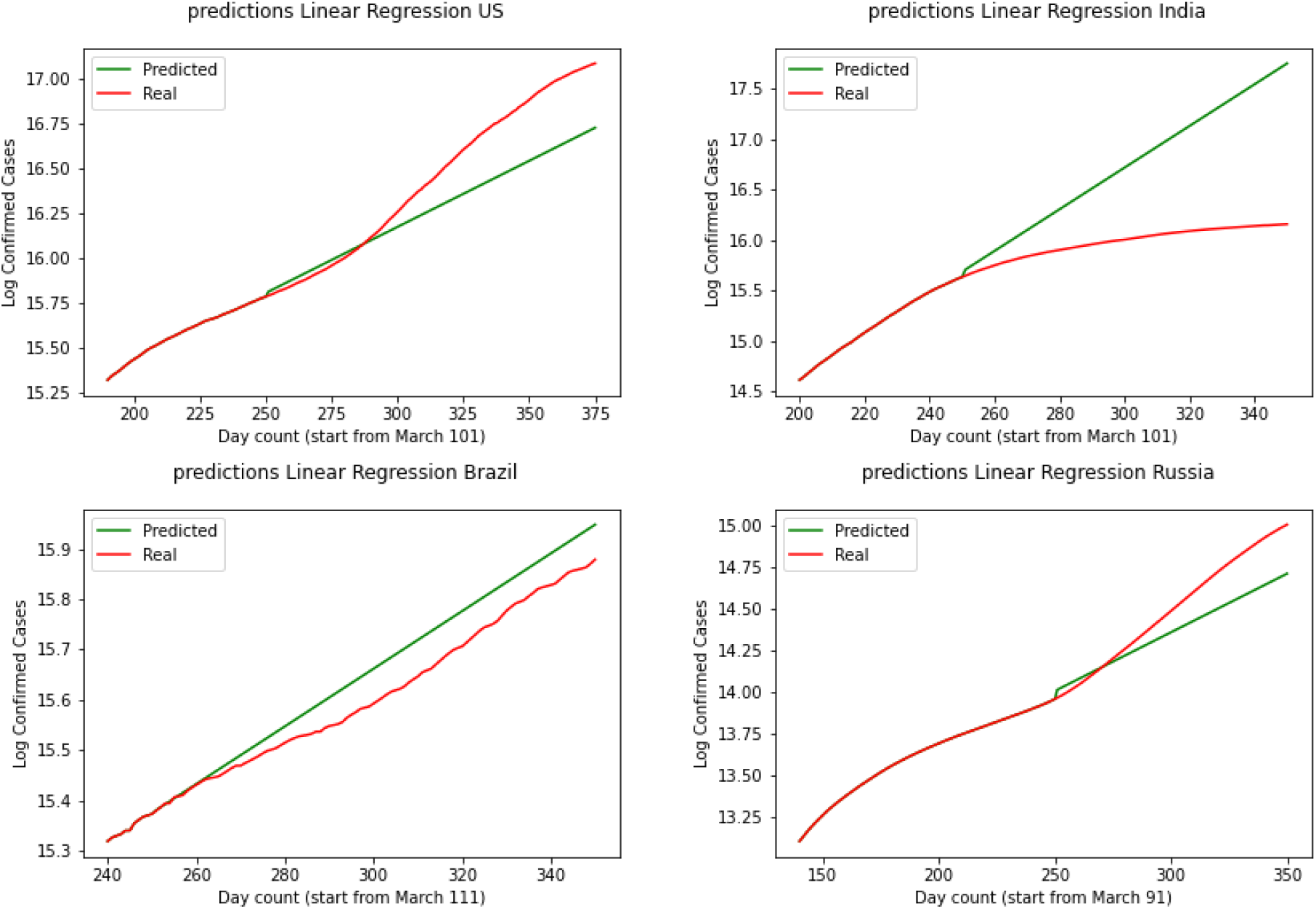
Linear regression

Where

*x*_0_ = sigmoid’s midpoint,

*L* = the curve’s maximum value,

**Fig. 7.**
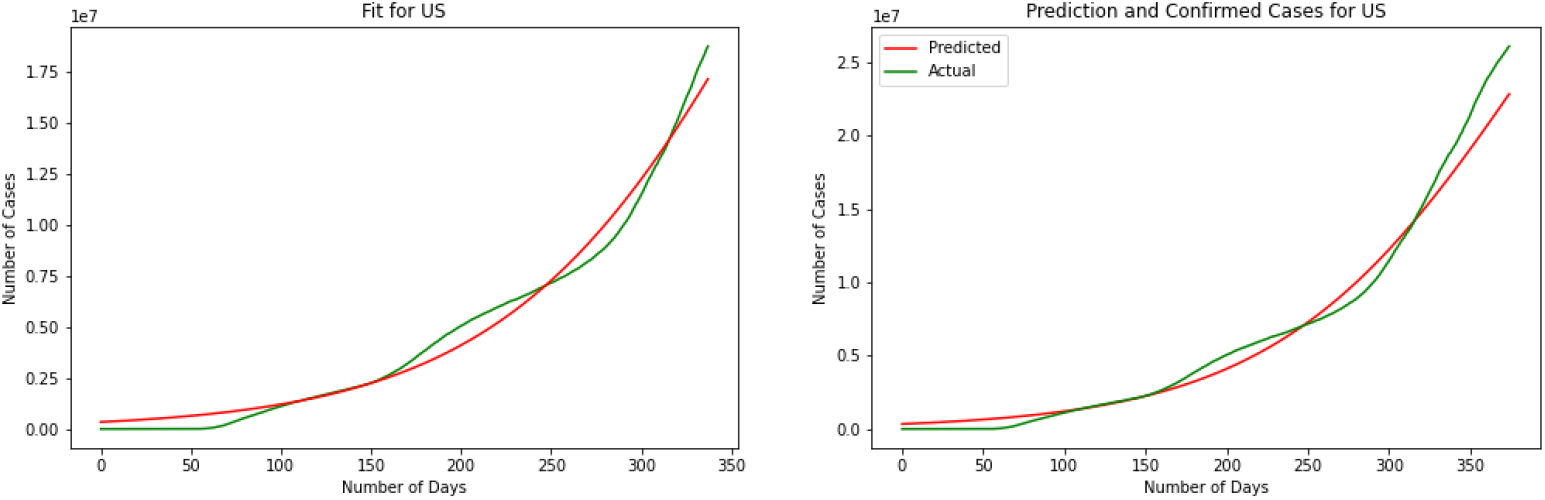
Logistic Curve for US

**Fig. 8.**
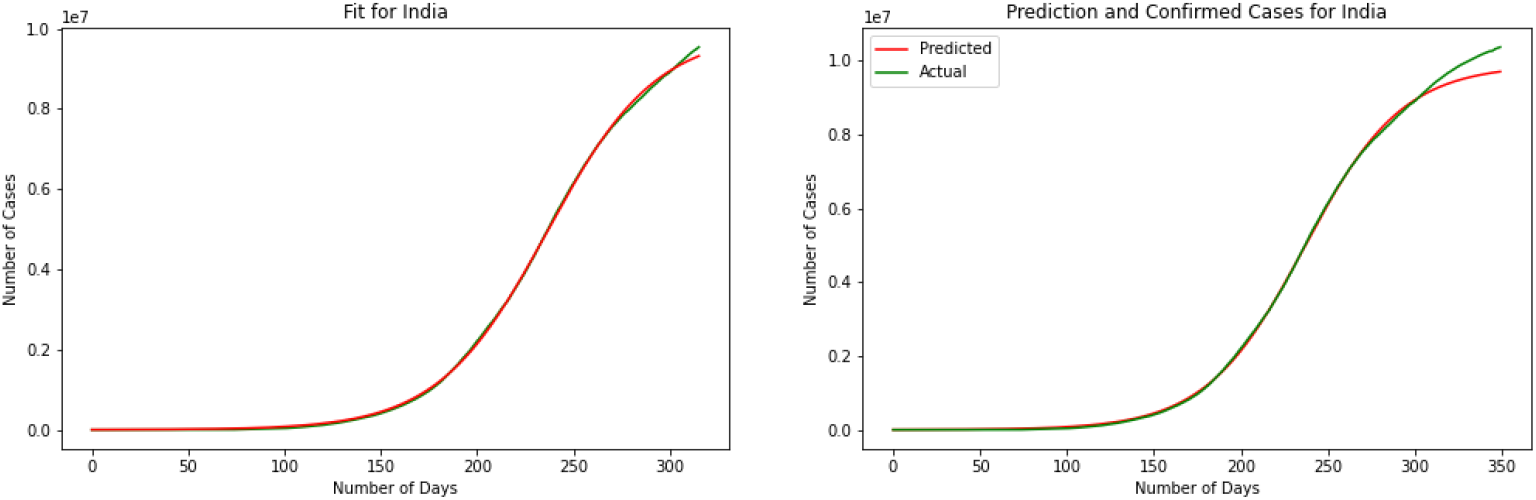
Logistic Curve for India

*k* = the logistic growth rate

This chart shows an examination of exponential and logistic growth with certain highlights featured. This equation communicates all the more plainly the essential propose underlying the logistic hypothesis, that the pace of growth diminishes linearly as the population increases. The underlying phase of growth is almost exponential ; at that point, as immersion starts, the growth eases back to linear, and at end, it stops the growth. The model can provide forecast for 3 out of 4 countries closely as the actual data. we have used 200 days to train the model and we have tested the model over the 50 days data after the 200 data. We have discovered a Logistic Function that is actually very near the watched Coronavirus information from theses four countries. As should be obvious in the diagrams beneath, the Logistic Model is truly not that a long way from the real nations Coronavirus information.

### 2.6 ARIMA Model

ARIMA model is a well known and generally utilized statistical technique for time series forecasting. ‘Auto Regressive Integrated Moving Average’ or ARIMA is a given time series dependent on its own previous values, in order to forecast future values using the equation. ‘non-seasonal’ time series with patterns which are not white noises can be modeled by ARIMA. ARIMA model was presented by Box and Jenkins in 1970. ARIMA models have demonstrated proficient ability to create short-term forecasts. ARIMA model is based on the idea that the variables future value is dependent on the past values of that variable and errors of that variable. It is a linear regression of the variable and the errors. This is conveyed as follows:

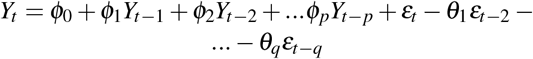

where,

*Y*_*t*_ is the real value,

*ε*_*t*_ is the random error at time t.

The steps in building ARIMA predictive model consist of model identification, parameter estimation and diagnostic checking.[23]. ARIMA model have been fitted to the data using 180 days as train data and the rest as test data. We can interpret from the charts that the model can be utilized in short term predictions since the data is changing in long term.

## 3 Conclusion

It is necessary to collect and analyze data of a pandemic to assess strategies of intervention, management, and control. This analysis gives a crucial baseline of the characteristics of the transmission and severity of the infectious disease. This study analyzes the behavior of the COVID-19 pandemic in United States. Moreover, a SIR-based model is presented in order to predict the number of cases and fatalities of this pandemic. Future research could use other models such as variations to the basic SIR model or individual-based network models. Comparisons among these models, in terms of accuracy and magnitude of error, could be made.

There are some extensions to the sir model that could be considered for the further studies. For example Satsuma has present a new discretisation of the SIR model which has the advantage of possessing a conserved quantity, thus making possible the estimation of the non-infected population at the end of the epidemic.[19]

**Fig. 9.**
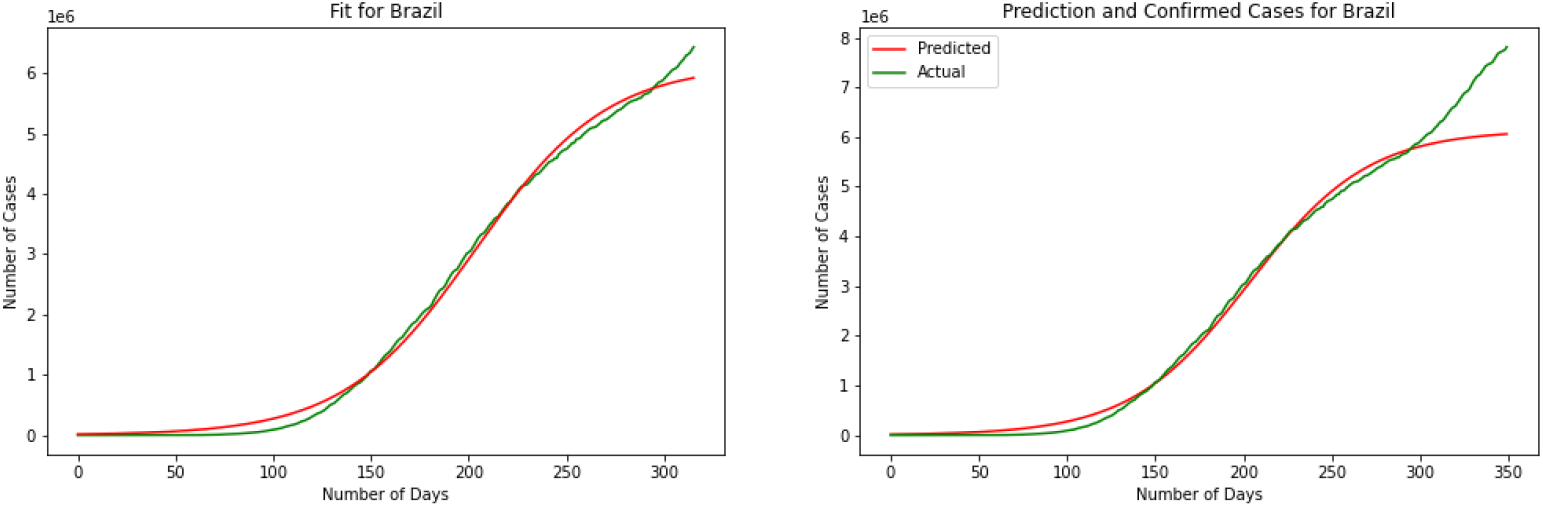
Logistic Curve for Brazil

**Fig. 10.**
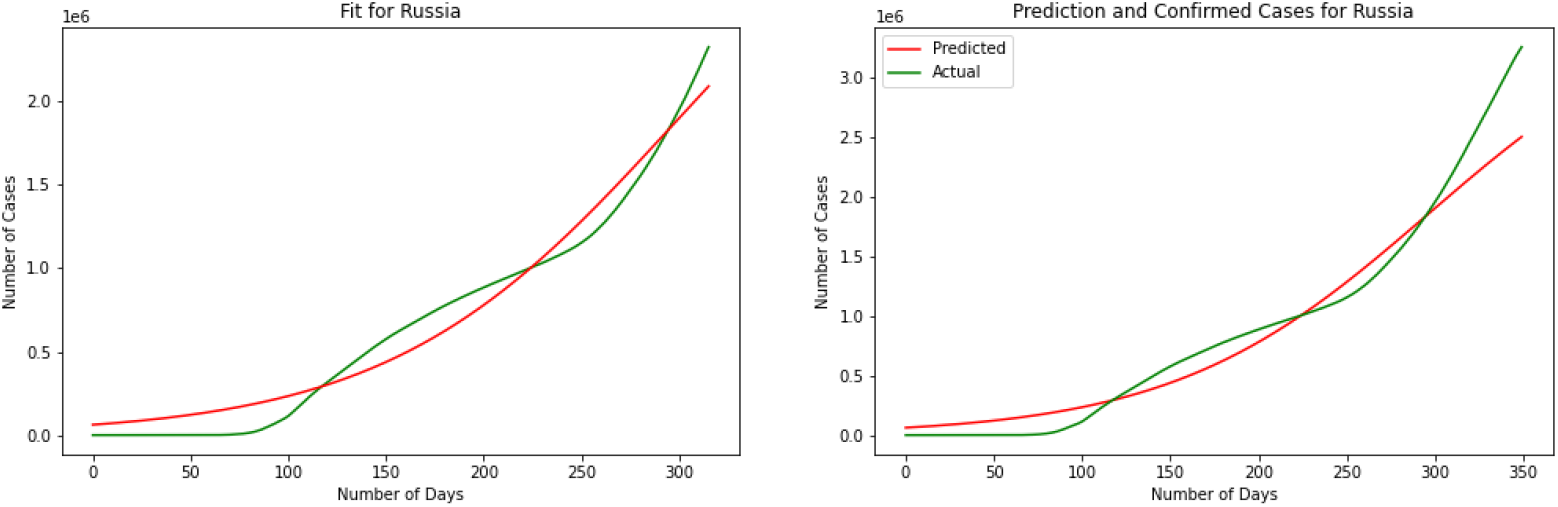
Logistic Curve for Russia

**Fig. 11.**
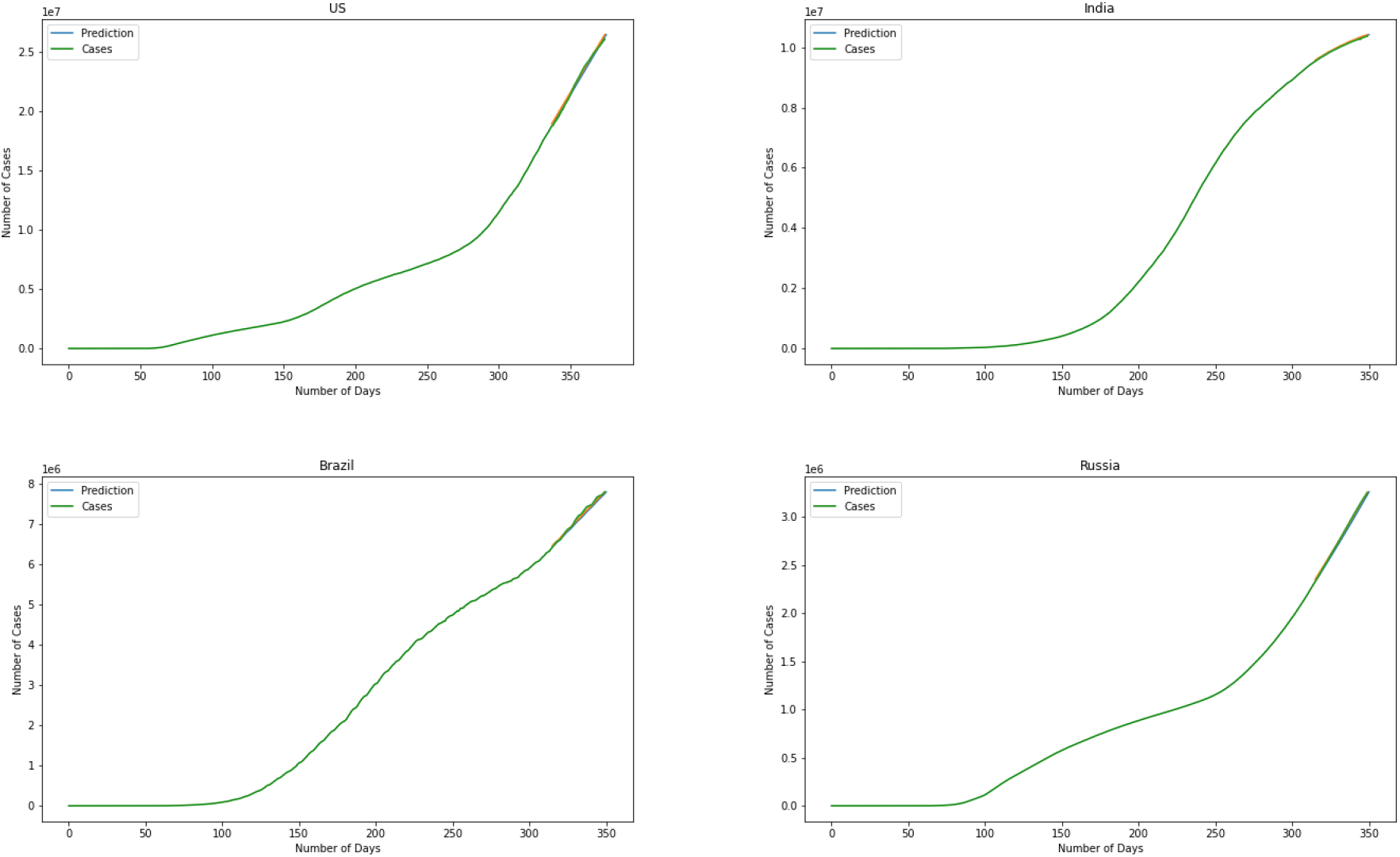
ARIMA Model

## Data Availability

or this research we have used GitHub data repository managed by Johns Hopkins University which contains daily time series summary tables, including confirmed, deaths and cases infected for more than once per day. Daily data of the influenced individuals are very helpful for data scientists. All data are from the daily case report, retrieved from: https://github.com/CSSEGISandData/COVID-19.

## Footnotes

1 Part of model was retrieved from the book Learning Scientific Programming with Python[21] and was coded in R

